# Seizure onset patterns predict outcome after stereotactic electroencephalography-guided laser amygdalohippocampotomy

**DOI:** 10.1101/2022.11.15.22282289

**Authors:** Andrew J. Michalak, Adam Greenblatt, Shasha Wu, Steven Tobochnik, Hina Dave, Ramya Raghupathi, Yasar T. Esengul, Antonio Guerra, James X. Tao, Naoum P. Issa, Garth R. Cosgrove, Bradley Lega, Peter Warnke, H. Isaac Chen, Timothy Lucas, Sameer A. Sheth, Garrett P. Banks, Churl-Su Kwon, Neil Feldstein, Brett Youngerman, Guy McKhann, Kathryn A. Davis, Catherine Schevon

## Abstract

**Objective:** Stereotactic laser amygdalohippocampotomy (SLAH) is an appealing option for patients with temporal lobe epilepsy, who often require intracranial monitoring to confirm mesial temporal seizure onset. However, given limited spatial sampling, it is possible that stereotactic electroencephalography (sEEG) may miss seizure onset elsewhere. We hypothesized that sEEG seizure onset patterns (SOPs) may differentiate between primary onset and secondary spread and predict postoperative seizure control. In this study, we characterized the two-year outcomes of patients who underwent single-probe SLAH after sEEG and evaluated whether sEEG SOPs predict postoperative seizure freedom.

**Methods:** This retrospective five-center study included patients with or without mesial temporal sclerosis (MTS) who underwent sEEG followed by single probe SLAH between August 2014 and January 2022. Patients with causative hippocampal lesions apart from MTS or for whom the SLAH was considered palliative were excluded. A SOP catalogue was developed based on literature review. The dominant pattern for each patient was used for survival analysis. The primary outcome was two-year Engel I classification or recurrent seizures before then, stratified by SOP category.

**Results:** 58 patients were included with a mean follow-up duration of 39 ± 12 months after SLAH. Overall one-, two, and three-year Engel I seizure freedom probability was 54%, 36%, and 33% respectively. Patients with SOPs including low voltage fast activity or low frequency repetitive spiking had a 46% two-year seizure freedom probability, compared to 0% for patients with alpha or theta frequency repetitive spiking or theta or delta frequency rhythmic slowing (log rank test, p = 0.00015).

**Significance:** Patients who underwent SLAH after sEEG had a low probability of seizure freedom at two years, but SOPs successfully predicted seizure recurrence in a subset of patients. This study provides proof of concept that SOPs distinguish between seizure onset and spread and supports using SOPs to improve selection of SLAH candidates.

**Key Points:**

- We described extended seizure outcomes in a five-center retrospective review of 58 patients.
- Seizure onset patterns (SOP) were categorized as putative positive vs. negative predictors of postoperative seizure freedom.
- Low voltage fast activity or low frequency repetitive spiking are associated with higher seizure freedom probability
- A 0% Engel I probability was found for patients whose dominant SOP was rhythmic slowing or repetitive spiking in the theta or alpha frequency bands.

## 1. Introduction

While anterior temporal lobectomy (ATL) has been the standard surgical treatment for patients with pharmacoresistant mesial temporal lobe epilepsy (mTLE), ^1^ stereotactic laser amygdalohippocampotomy (SLAH) is a newer, less invasive alternative with lower complication rates and possibly fewer neuropsychiatric side effects.^2,3^ However, although seizure freedom rates are comparable to ATL in the first year after SLAH, the probability of seizure freedom falls below 50% beyond two years.^4^ Moreover, it is unclear to what degree patients requiring stereotactic electroencephalography (sEEG) to identify the seizure focus may benefit from SLAH as these patients often have clinical and imaging features discordant with mTLE. One study of 12 patients without mesial temporal sclerosis (MTS) reported that seven attained Engel I seizure freedom after SLAH if sEEG confirmed mesial temporal seizure onset.^5^ However, only one of the seizure free patients had at least two years of follow-up. A recent meta-analysis of laser interstitial thermal therapy (LITT) showed a 42% prevalence of seizure freedom in 69 patients across five studies examining LITT after intracranial EEG, compared to the overall prevalence of 56% with or without intracranial EEG. Of note, the majority of the 69 patients underwent LITT of non-mesial temporal structures.^6^ Thus, the long-term efficacy of mesial temporal ablation after sEEG is not well-characterized.

The interpretation of hippocampal seizure onset with sEEG is not necessarily straightforward. Since seizure onset, propagation, and semiology can happen rapidly, distinguishing primary hippocampal onset from secondary propagation can be challenging. This is especially important in considering SLAH since seizure onset may occur in anterior temporal neocortex or parahippocampal gyrus (PHG) which may not be adequately sampled in a sEEG array. These structures are resected in ATL but are spared in SLAH. Furthermore, prior studies show that ictal activity seen on electrocorticography, while generally interpreted to indicate seizure onset, may instead represent a local response to a seizure generated elsewhere.^7-11^Prior literature supports using intracranial EEG visual seizure onset pattern (SOP) morphology for predicting epilepsy surgery outcomes. For instance, certain SOPs have been associated with areas of secondary propagation rather than primary onset.^12^ However, many studies on the prognostic value of SOPs were in the context of ATL.^13,14^ Since ATL removes the amygdalohippocampal complex (AHC) as well as the PHG, temporal pole, and lateral temporal neocortex, it is unclear whether SOPs that portend a favorable prognosis in ATL are applicable to SLAH. At the same time, due to its spatial specificity and sparing of the PHG and surrounding neocortex, SLAH offers an ideal proof of concept for studying the predictive value of sEEG biomarkers.

Few studies exist leveraging SOPs with focal ablations. One series of 35 patients showed that sEEG SOPs are helpful in predicting good outcome after laser interstitial thermal therapy for extra-temporal lobe epilepsy.^15^ Another series of nine patients with mTLE who underwent laser ablation after sEEG demonstrated that frequencies below 14 Hz, regardless of SOP, at hippocampal seizure onset were associated with a worse prognosis compared to frequencies above 14 Hz.^16^In this study, we present a survival analysis for sEEG-guided SLAH and explore factors associated with two-year seizure freedom. We then generated a catalogue of SOPs based on prior literature and analyzed patient outcomes based on their sEEG SOP.

## 2. Materials and Methods

### 2.1 Patient selection and stereo EEG recordings

We performed a five-center retrospective analysis of consecutive patients with mTLE treated with sEEG-guided single-probe SLAH between August 2014 to January 2022 (specific dates for each center varied within this time window). The sites involved were Columbia University, University of Chicago, University of Pennsylvania, Brigham and Women’s Hospital, and University of Texas-Southwestern. Retrospective patient data collection and analysis was conducted in accordance with protocols approved by each center’s Institutional Review Board.

Patients with pharmacoresistant epilepsy underwent presurgical evaluation according to standard clinical practice at their respective institutions. Such patients underwent intracranial implantation due to discordant data, or high suspicion of a neocortical seizure focus based on the presurgical evaluation. Patients were included with or without magnetic resonance imaging (MRI) evidence of MTS,^17^ or abnormalities thought to be unrelated to their seizures based on presurgical and sEEG findings (e.g., small cavernous malformations distant to the theorized epileptogenic zone). Patients were excluded if they had a lesion other than MTS thought to be driving their seizures (e.g., glioma or focal cortical dysplasia visualized on MRI). Patients were also excluded if SLAH was considered palliative (e.g., multiple seizure onset zones identified but SLAH was performed to decrease the frequency of a single seizure type), if they had multiple ablation targets, or if the approach utilized two probes.

All patients were evaluated using sEEG depth arrays with 8-16 contacts each. The electrode manufacturer, specifications, recording and reviewing equipment were institution specific. One center (University of Chicago) used subdural strips together with depth arrays in a subset of patients. Patients were excluded if they demonstrated onset in the strip contacts prior to onset in the depth contacts. Electrode positioning in the hippocampus and amygdala were verified by co-registration of the post-implant CT with pre-operative MRI at the time of implant and confirmed by review of records at each institution.

### 2.2 Seizure onset pattern catalogue

A catalogue of nine seizure onset patterns (SOPs) was developed based on analysis of current literature^16,18-22^ and our observations from microelectrode recordings (Figure 1).^7,11,23,24^ Each SOP was classified based on expected prognosis for seizure outcome (Table 1). A SOP was categorized as a “favorable pattern” if prior literature showed a higher likelihood of seizure freedom following open resection, or if the pattern was associated with primary seizure onset. Likewise, a pattern was categorized as an “unfavorable pattern” if prior literature showed a low likelihood of seizure free outcomes following open resection, or if the pattern was consistently associated with areas of secondary seizure propagation. {Table 1 about here}

**Figure 1.**
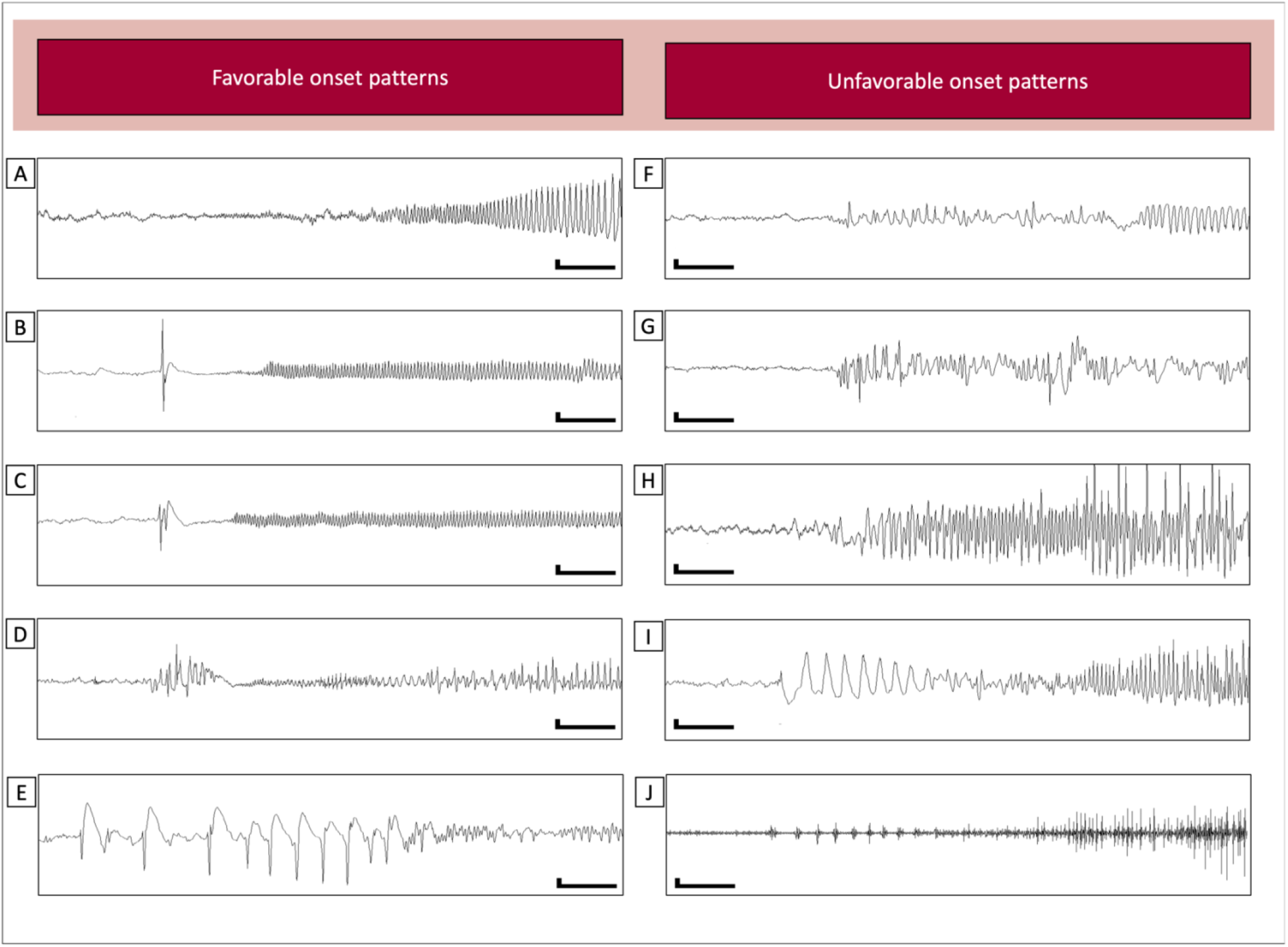
The nine seizure onset patterns broken down by association with a favorable prognosis (A-E) or unfavorable prognosis (F-I). A. Low voltage fast activity (LVFA). B. Herald spike followed by LVFA (HS>LVFA). C. Herald polyspike followed by LVFA (HPS>LVFA). D. Herald burst followed by LVFA (HB>LVFA). E. Low frequency repetitive spiking. F. Repetitive spiking in the theta frequency range (RSPT). G. Repetitive spiking in the alpha frequency range (RSPA). H. Repetitive spiking in the beta frequency range (RSPB). I. Rhythmic slowing in the delta frequency range. J. Superimposed rhythmic fast activity (“brush”) overriding the delta slowing depicted in H. A-I: highpass filter: 1Hz, lowpass filter: off, scale legend 300uV/1 second; I: highpass filter: 50, lowpass filter: 250, scale legend 140uV, 1 second.

**Table 1.** Seizure onset pattern catalogue.

| <u>Seizure onset patterns associated with a favorable prognosis</u> |  |  |
| --- | --- | --- |
| Name | Abbreviation | Description |
| Low voltage fast activity | LVFA | Rhythmic activity > 13 Hz, usually < 10uV in initial amplitude with gradually increasing amplitude |
| Herald spike / polyspike / burst followed by LVFA | HS>LVFA<br>HPS>LVFA<br>HB>LVFA | A single spike, polyspiking, or spike bursts lasting < 5 seconds, followed by LVFA |
| Low frequency repetitive spiking | LFRS | ≤ 3 Hz, high amplitude, lasting for > 5 seconds |
| <u>Seizure onset patterns associated with an unfavorable prognosis</u> |  |  |
| Name | Abbreviation | Description |
| Rhythmic theta/delta slowing or delta brush | RSL | Rhythmic onset of delta or theta sinusoidal/slowing activity, with or without embedded high frequency activity ("brush") |
| Repetitive spiking in the theta, alpha, or beta frequency band | RSPT, RSPA, RSPB | High amplitude rhythmic spikes between 5-25 Hz frequency band with, lasting for > 5 seconds |

#### Patterns associated with a favorable prognosis

1. *Low-voltage fast activity (LVFA)*. Fast (>13 Hz) oscillatory activity generally <30-50 uV at onset. This pattern is widely described and associated with a favorable surgical outcome in neocortical and mesial temporal epilepsy.^20^
2. *Herald spike, polyspike, and burst followed by low-voltage fast activity (HS>LVFA, HPS>LVFA, HB>LVFA)*. In vivo neocortical multiunit recordings suggest that local seizure onset may be associated with LVFA when *preceded* by a herald spike.^7^ Additionally, the onset pattern of polyspikes lasting < 5 seconds and followed by LVFA was associated with focal cortical dysplasia and a favorable surgical,^22^ but this pattern has not previously been reported in hippocampal seizures. To explore the prevalence and relevance of these patterns in hippocampal seizures, we defined them as a single spike, polyspike complex, or burst(s) of polyspiking lasting <5 seconds and immediately followed by the onset of LVFA. To classify as a herald pattern, the spikes must be distinct from the interictal background, and multiple seizures may be evaluated to determine whether the heralding spike is separate from background interictal populations.
3. Low frequency repetitive spiking (LFRS). High amplitude spiking ≤ 3 Hz and lasting for > 5 seconds, which has been referred to as “preictal spiking”^18,25^ and widely reported. This pattern is associated with hippocampal neuronal loss and mesial temporal sclerosis, and with a favorable surgical prognosis.^20,22^

#### Patterns associated with an unfavorable prognosis

1. *Repetitive spiking in the theta, alpha, or beta frequency range (RSPT, RSPA, RSPB)*. High amplitude repetitive spikes in the theta, alpha, or beta frequency range (5-25 Hz), lasting for > 5 seconds. Repetitive spiking at various frequencies has been associated with better or worse surgical outcomes, and there is considerable heterogeneity of which frequency ranges are included in analyses.^20^ Prior studies in patients undergoing resection^21,26^ and one study in SLAH^16^ showed that alpha or theta frequency activity is associated with a worse prognosis than high frequency activity.
2. *Rhythmic slowing and delta brush (RSL)*. Rhythmic slowing in the theta or delta frequency range, with or without overriding high frequency activity (i.e. “brush”). These patterns have been associated with secondary seizure propagation^12^ and a worse surgical outcome.^22^

#### ectrodecrement

In addition to the above patterns, we added electrodecrement (ED) as a modifier and defined it as attenuation of background amplitude with or without superimposed fast activity.^27^ ED was categorized as focal if it occurred only in the electrodes demonstrating maximal seizure onset or diffuse if it extended to electrodes outside of the temporal lobe that were not part of the primary seizure onset (Figure 2).

**Figure 2.**
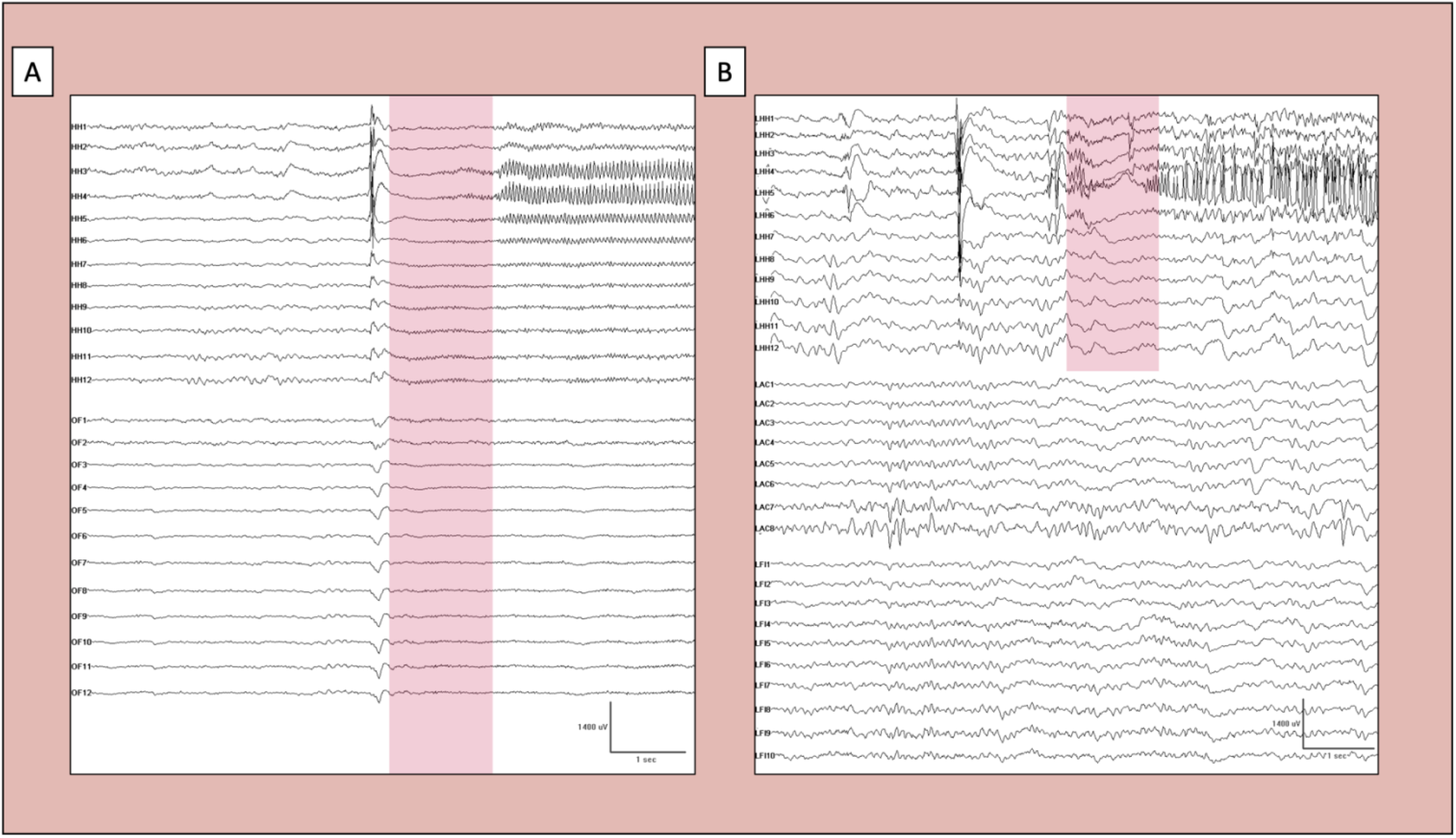
Electrodecrement (shaded areas). A. Diffuse electrodecrement which manifests as attenuation with or without superimposed faster frequencies at the site of seizure onset (hippocampal head; HH) as well as the lateral contacts and more distant contacts that were not primarily involved with the seizure (orbitofrontal cortex; OF). B. Focal electrodecrement constrained to the site of seizure onset (hippocampal head—LHH) but not prominently in the lateral or extratemporal contacts. Note the ongoing oscillatory activity in the anterior cingulate (LAC) and frontal to insular (LFI) contacts.

### 2.3 Seizure onset pattern scorings

Seizure onset patterns (SOPs) were analyzed independently by two epileptologists blinded to clinical outcomes (A.J.M. at all sites; C.A.S. at Columbia University; A.G. at University of Pennsylvania; S.W. at University of Chicago; S.T. at Brigham and Women’s; H.D. at University of Texas Southwestern). All seizures were evaluated by the primary site reviewer. Representative examples of each patient’s SOPs as well as ambiguous SOPs were secondarily reviewed by A.J.M. (C.A.S. at Columbia). Seizure onset was defined as the first clear departure from the interictal background, and the SOP was graded on the first pattern seen. If a consensus could not be reached by both reviewers, the seizures were classified by consensus of two senior epileptologists (C.A.S. and K.D.), again blinded to outcome. Reviewers were allowed to use multiple montages and bandpass filters to reach a decision. If a patient had multiple SOPs seen across seizures, the dominant pattern (most frequently occurring pattern) was used for analysis.

### 2.4 Laser ablation

Standard single-probe SLAH was performed as described elsewhere^2^ and entailed stereotactic placement of a laser fiber and MRI thermometry during the ablation. Each institution performed the procedure in accordance with its own practices. Operative notes were reviewed to ensure each procedure was in accordance with the traditionally described approach, and deviations were noted (e.g., limiting the trajectory due to atypical anatomy). Authors at each center reviewed postoperative imaging to confirm that the ablation cavity included the intended targets, i.e. amygdala and hippocampal head and body.

### 2.5 Clinical variables

Patient information collected included clinical history, MRI and positron emission tomography (PET) findings, epilepsy risk factors, and seizure types. Surgical outcome was based on review of postoperative follow-up records and rated in accordance with the Engel classification scale.^28^ Seizure freedom was defined as an Engel I outcome (i.e. freedom from disabling seizures), and failure was defined as an Engel II-IV outcome. Patients with any duration of follow-up were included for survival analysis. The selection of two-year outcomes for SOP analysis was based on our observation, consistent with prior studies, that seizure freedom after SLAH drops sharply through the first two years postoperatively. As such, patients who were seizure free but with less than two years of follow-up were excluded from subgroup analysis.

### 2.6 Statistical analysis

Descriptive statistics were presented as mean (range) and percentages. Kaplan-Meier survival curves were generated from the overall cohort and dichotomized by favorable vs. unfavorable prognostic pattern. Comparison of survival curves for onset patterns was performed using the log-rank test. Fisher’s two-tailed exact test was used for subgroup analysis. A P ≤ 0.05 was considered statistically significant. Statistical tests and figures were generated with Python 3.9.7 using publicly available packages.^29-31^

## 3. Results

### 3.1 Population

Sixty patients were identified and 58 were included for analysis (35 female; mean age 39 years; range 20-66 years; Table 2). Two patients were excluded in whom seizure outcome could not be graded: one with epileptic encephalopathy without discrete electroclinical seizures and another with a profound nonepileptic seizure burden. The mean follow-up duration was 39 (± 26) months. MTS was seen in 14 patients and was more common in patients with favorable SOPs (13 patients with favorable SOPs had MTS versus only one patient with an unfavorable SOP).

**Table 2.** Patient Characteristics. Due to the nature of the ablation procedure, tissue pathology was not available.

| **Table 2. Patient Characteristics** |  |  |  |
| --- | --- | --- | --- |
|  | **Favorable SOPs** | **Unfavorable SOPs** | **Total** |
| N | 43 | 15 | 58 |
| Sex, female (%) | 26 (60) | 9 (60) | 35 (60) |
| Age in years, mean (range) | 40 (20-66) | 37 (21-54) | 39 (20-66) |
| Age of onset of epilepsy in years, mean (range) | 25 (0-52) | 24 (1-47) | 25 (0-52) |
| Follow up in months, mean (range) | 40 (3-87) | 35 (6-82) | 39 (3-87) |
| **Laser side** |  |  |  |
| Right | 20 (47) | 9 (60) | 29 (50) |
| **Implant side** |  |  |  |
| Bilateral (%) | 21 (49) | 3 (20) | 24 (41) |
| Right (%) | 11 (26) | 8 (53) | 19 (33) |
| Left (%) | 11 (26) | 4 (27) | 15 (26) |
| **Risk factors** |  |  |  |
| Febrile seizure (%) | 5 (12) | 3 (20) | 8 (14) |
| Family history (%) | 6 (14) | 1 (7) | 7 (12) |
| Preterm/abnormal pregnancy (%) | 3 (7) | 2 (13) | 5 (9) |
| Head trauma (any severity) (%) | 8 (19) | 2 (13) | 10 (67) |
| CNS infection (%) | 1 (2) | 2 (13) | 3 (5) |
| Hemorrhage (%) | 1 (2) | 0 (0) | 1 (2) |
| Autoimmune (%) | 1 (GAD65+) (2) | 0 (0) | 1 (2) |
| No risk factors (%) | 21 (49) | 7 (47) | 28 (48) |
| **Seizure type** |  |  |  |
| Focal aware seizure (%) | 18 (42) | 6 (40) | 24 (41) |
| Focal impaired aware seizure (%) | 39 (91) | 15 (100) | 54 (93) |
| Secondary generalization (%) | 39 (91) | 12 (80) | 51 (88) |
| **Imaging** |  |  |  |
| MTS | 13 (30) | 1 (7) | 14 (24) |
| PET concordant | 31 (72) | 10 (67) | 41 (71) |
| PET not concordant | 11 (26) | 5 (33) | 16 (28) |
| PET normal | 3 (7) | 3 (20) | 6 (10) |
| Bilateral PET abnormalities | 5 (12) | 1 (7) | 6 (10) |
| Extratemporal PET abnormalities | 3 (7) | 1 (7) | 4 (7) |

By two years, 48 patients had either failed or were seizure free with follow-up data and were included in year-two analysis. Review by senior epileptologists was required in one case where there was ambiguity between repetitive spiking (unfavorable SOP) and herald bursts (favorable SOP). This was due to the varying length of spiking across seizures (in some seizures lasting > 5 seconds and others < 5 seconds. The consensus was to grade this as herald bursting, since the spiking was usually less than 5 seconds. {Table 2 about here}

The probability of seizure freedom in the cohort was 54%, 36%, and 33% at one, two, and three years, respectively (figure 3a). There was no difference in two-year seizure freedom for patients with MTS, with concordant PET positivity, or when combining patients with MTS and concordant PET positivity (all p > 0.1; Figure 3B). Likewise, when excluding all patients with MTS, patients with favorable SOPs still had a higher probability of seizure freedom (p = 0.0016, log rank).

**Figure 3.**
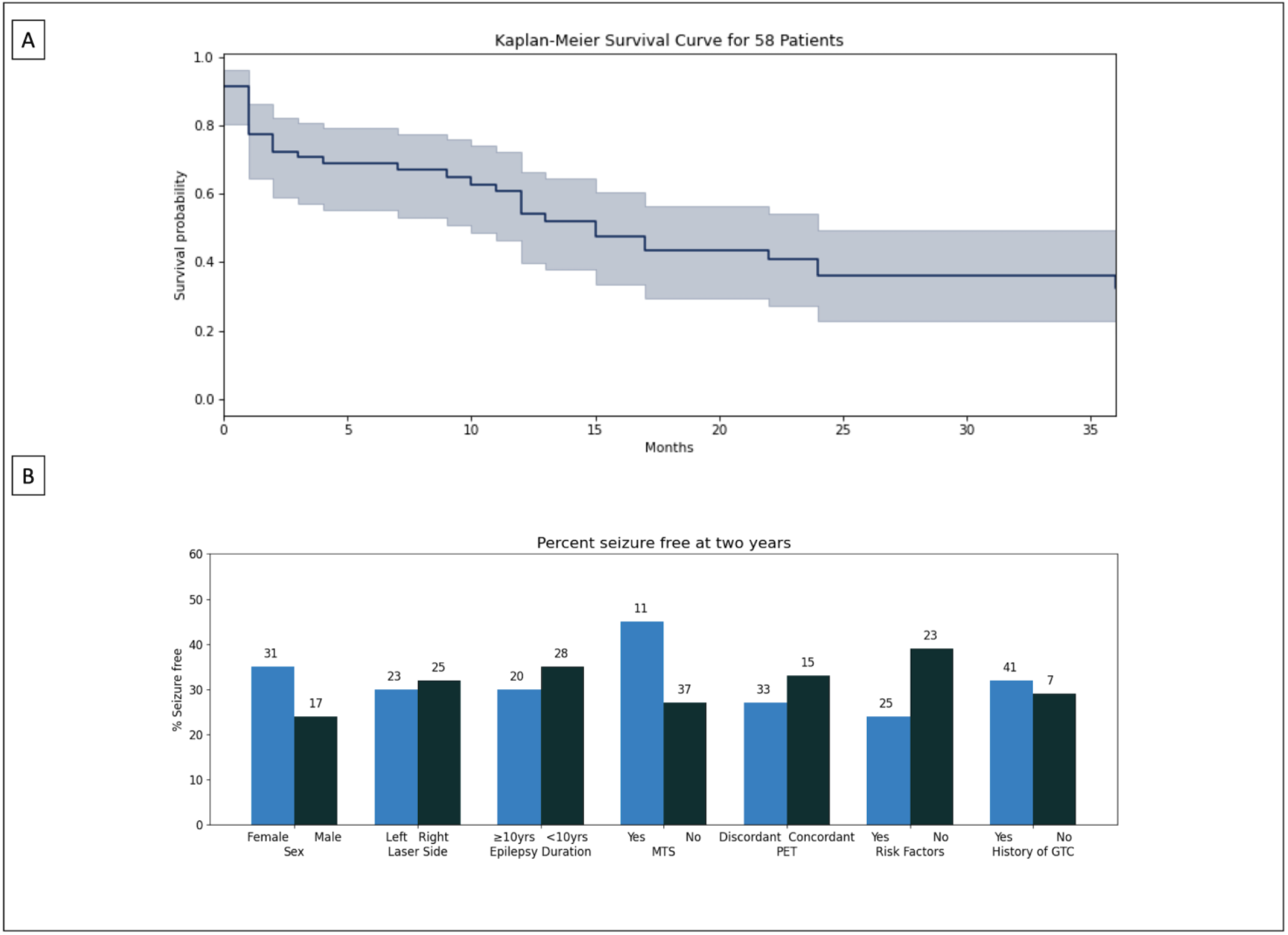
A. Kaplan-Meier curve depicting Engel I outcomes for entire surgical cohort. At one year the Engel I probability was 54%. This fell to 36% by year 2 and remained stable (33%) at year 3. This trend informed our decision to limit the SOP predictor study to 2-year outcomes. Shaded area represents 95% confidence intervals. B. Two-year seizure freedom in 48 patients with either >=2 years of follow-up or failure prior to 2 years. Absolute number of seizure free patients for each subgroup is stated above each bar. No variables reached significance on individual testing.

### 3.2 Seizure onset pattern prevalence

LVFA with or without preceding transient features comprised 55% of all dominant patterns (LVFA: 20.7%, HS>LVFA: 6.9%, HPS> LVFA: 12.1%, and HB>LVFA: 15.5% (figure 4a).

**Figure 4.**
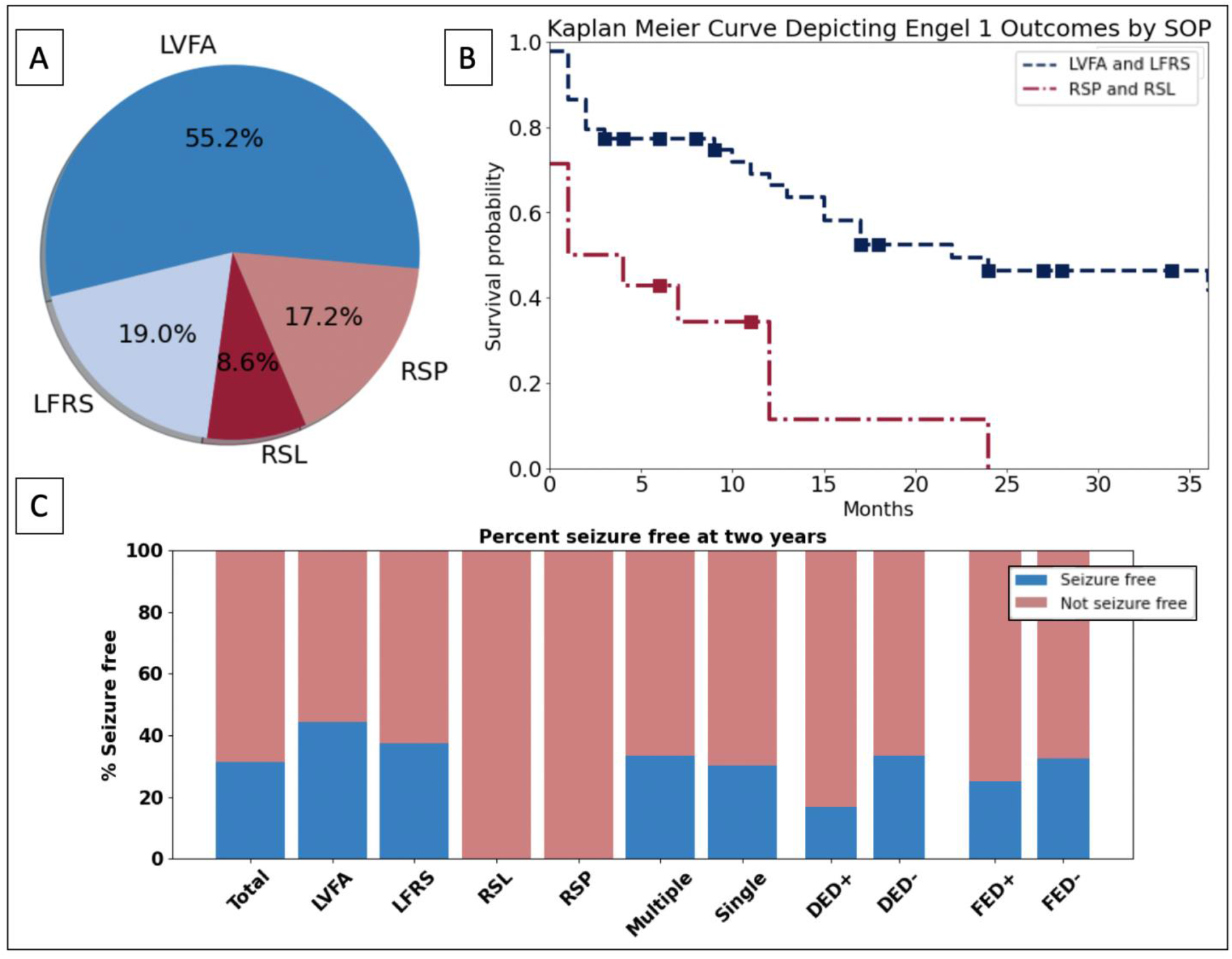
A. Breakdown of dominant pattern occurrence. LVFA-containing SOPs were the dominant pattern in 55.2% of patients, followed by LFRS. B. Kaplan-Meier curve depicting Engel I outcomes dichotomized by seizure onset pattern. Patients with a dominant SOP containing LVFA or LFRS had a one-year seizure freedom probability of 66% compared to 11% for patients with RSPA, RSPT, or RSL. At two years, the survival probability was 46% and 0%, respectively (p = 0.00015). C. Two-year seizure freedom broken down by individual SOPs. SOPs containing LVFA or low frequency repetitive spiking had better outcomes at two years when compared to rhythmic slowing or repetitive spiking in the theta or alpha frequency band. Outcomes did not differ if the patient had a single SOP vs. multiple SOPs, or if the patient’s seizures demonstrated diffuse or focal electrodecrement (DED and FED, respectively).

Low frequency repetitive spiking comprised 19% of dominant patterns. Theta and alpha frequency spiking was seen in 17.2% of patterns. Beta frequency spiking was not observed as a dominant pattern. Multiple SOPs, i.e., SOPs varying across seizures recorded from an individual, were observed in 16 patients (28%). There was no predilection for which patterns co-occurred.

Focal ED was noted in the seizures of ten patients and diffuse ED was seen in seven patients. Focal ED was seen mostly in the context of LVFA-containing SOPs (three LVFA, two HB>LVFA, two HPS>LVFA, one HS>LVFA) and once in RSPT. Diffuse ED was seen in HPS>LVFA (4), and once each in LFRS, RSPT, and RSL.

### 3.3 Seizure onset patterns predict post-ablation outcome

Patients with favorable SOPs had a 46% probability of two-year seizure freedom, compared to a 0% probability for patients with unfavorable SOPs (p = 0.00015; figure 4b). The median time to seizure recurrence for patients with favorable SOPs was 9.5 months, compared to 1 month for patients with unfavorable SOPs. All but one patient with unfavorable SOPs had recurrent seizures in the first year.

#### SOP-specific outcomes

Twelve of 27 patients with LVFA (44%) and 3 of 9 patients with LFRS (33%) were seizure free at two years. LFRS was not associated with MTS (5/12 patients with MTS had LFRS and 9/46 without MTS had LFRS; p = 0.3). None of the twelve patients who demonstrated RSPT/RSPA or RSL as a dominant SOP were seizure free at two years (figure 4c).

Compared to patients with a single SOP, patients who had multiple SOPs had similar outcomes at two years (10/33 seizure free in the single SOP group vs 5/15 in the multiple SOP group; p >0.05). Though this study was not powered to directly compare these subgroups, we observed that all nine patients who had RSL either as a dominant pattern or as one of multiple patterns had seizure recurrence.

Overall ED outcomes were 3/14 (21%) seizure free at two years (two of eight with focal ED and one of six with diffuse ED), compared to 13/34 (38%) without ED. When restricted to patients who demonstrated SOPs with a putative favorable prognosis, three of ten patients with ED (30%) were seizure free at two years.

### 3.4 Effects of deviations in ablation trajectories

Seven patients had slight deviations in the ablation trajectory or lesion on review of postoperative imaging. Two patients had some residual amygdala on postoperative MRI; three patients had slightly altered fiber trajectories (two inferior and one superior), and two had modifications to their ablation target (one targeting the hippocampal body, and the other with a standard trajectory but with incomplete ablation of the posterior hippocampal body). The results of the primary analysis were not changed after excluding these patients (p = 0.00044).

## 4. Discussion

Although SLAH is a less invasive and lower morbidity alternative to ATL for many patients with pharmacoresistant epilepsy,^2,3^ little is known about how it performs in patients with discordant data requiring sEEG evaluation to localize the seizure onset zone. In this study, we demonstrated one-through three-year outcomes in patients undergoing SLAH after sEEG. Then, using a SOP catalogue, we dichotomized these SOPs into favorable or unfavorable pattern categories and identified a subset who have a high probability of failure within two years.

One-, two-, and three-year seizure freedom probability in our cohort was 54%, 36%, and 33%, respectively (figure 3). The drop between one- and two-year outcomes is consistent with prior literature^4^ and cautions against using one-year seizure freedom to study prognostic factors related to SLAH.

The overall outcomes in this cohort are poor compared to larger SLAH studies that include patients with or without preceding sEEG^4^. By limiting our study to patients who underwent sEEG, we selected a more complex patient population who could not be diagnosed with mTLE based solely on the noninvasive presurgical evaluation. Similar outcomes have been found in other sEEG-guided ablation cohorts (see Barot 2022 for review), though few of these have focused on laser ablation for mTLE.^5,32^ Comparing these outcomes to historical outcomes in sEEG-guided ATL,^33,34^ these cohorts performed worse at two years. However, given the potential benefits of SLAH over larger open resection, identifying predictive biomarkers will help identify ideal candidates.

### 4.1 Clinical significance of individual SOPs

No patient demonstrating RSPT/RSPA was seizure free at two years. The presence of spiking in these frequency bands has been associated with worse outcome when compared to high frequency activity.^16,21,26^ Similarly, none of the nine patients demonstrating RSL (either as a dominant pattern or as one of multiple patterns) were seizure free at two years. Rhythmic slowing in the theta or delta frequency range has been associated with a poor prognosis across several studies. It is commonly seen in areas of seizure propagation,^11,12^ and in the context of the current study, may have represented cases in which seizure onset was elsewhere, but captured on sEEG after hippocampal spread. These findings suggest that RSPT/RSPA and RSL serve as reliable biomarkers of poor response to SLAH.

Consistent with several other studies in both neocortical and mesial temporal epilepsy,^15,20^ LVFA was associated with a higher likelihood of postoperative seizure freedom. This finding was seen independently of whether the LVFA was preceded by spikes or bursts. The importance of a herald wave or waves preceding LVFA is not well-studied, in part because earlier analyses of SOPs excluded findings preceding the onset of sustained rhythmic activity. One study of 69 patients who underwent resection reported preictal discharges associated with seizure onset in 52 patients and found that its presence did not adversely affect resection outcome even when the discharge was bilateral or diffuse.^21^ Another analysis of the SOPs of 252 patients found herald bursting followed by LVFA in fifteen patients with FCD and noted that it was associated with a good post-resection prognosis.^22^

Concordant with previous studies,^20,22^ we found LFRS was associated with higher rates of seizure freedom. While we did not observe an association with mesial temporal sclerosis in contrast to prior studies,^13,25,35^ this could be due to inadequate power.

Twenty six percent of patients demonstrated multiple SOPs, but this did not affect seizure freedom. Although many prior studies either excluded patients with multiple SOPs or did not mention them,^20^ their role in outcomes remains unclear. Good outcomes after resective surgery range from 42%^36^ to 82%.^35^ Less is known about its importance in mesial temporal resections specifically, though a study examining single vs. multiple SOPs in patients who underwent resection of mesial temporal structures found that multiple SOPs were not associated with a worse outcome.^37^

ED^27^ was seen in fifteen patients (eight patients with focal ED; five patients with diffuse ED; two patients with both focal and diffuse ED). ED in sEEG is variably reported. Lagarde et al. did not observe diffuse ED in any of 820 seizures across 252 patients, positing that this may be a signature of grid recordings of a distant seizure onset.^22^ Conversely, Dolezalova et al.^19^ and Jiménez-Jiménez et al.^21^ found diffuse ED was associated with poor outcomes in patients undergoing sEEG. The variable reporting may reflect differences in definitions of ED and underscore the need for standardized definitions or quantitative analysis. Our study did not show worse outcomes in patients with ED, but this may be due to inadequate sample size to detect differences.

### 4.2 Mesial temporal sclerosis

We found no association between MTS and seizure freedom. The role that MTS plays in determining ablation outcomes is controversial. While a recent systematic review showed improved outcomes in MTS mTLE cases in SLAH whether the patient underwent sEEG or not,^6^ individual studies focusing on sEEG-guided SLAH have found differing results. Youngerman et al. found no association between MTS and outcome after SLAH,^5^ whereas Tao et al. found that patients with MTS were more likely to become seizure free.^32^ The lack of pathological diagnosis may result in misclassification of patients with MRI-negative MTS. Further, we found no association between concordant PET hypometabolism and seizure freedom, or when combining MTS and PET concordant cases. While MTS was more common in the favorable SOP group, the findings of the survival analysis were still significant when we excluded patients with MTS.

### 4.3 Limitations

There are several limitations to this study. Although we sought to strengthen our findings by conducting a multicenter study and defining SOPs *a priori*, the study was underpowered for detection of smaller subgroup effects in multivariable analysis, or to assess each SOP independently.

Surgical technique and ablation targets play a role in SLAH outcomes. While some studies did not find an association between ablation volume and seizure freedom,^38,39^ a larger multicenter study found improved seizure freedom with increased ablation volume of the amygdala and inclusion of the hippocampal head, PHG, and rhinal cortex.^40^ Another analysis of 28 patients found that patients with Engel I outcomes had a higher percentage of PHG ablated,^41^ and a recent study of 11 patients undergoing a two-trajectory approach to target the piriform and entorhinal cortex suggested improved outcomes.^42^ Conversely, other studies have found no correlation with ablation of these regions and outcome.^43,44^ Some centers now use two fibers including an oblique lateral entry point to cover the amygdala and entorhinal cortex. Although the results of our primary analysis did not change when we excluded patients who had an altered or incomplete ablation, we did not perform quantitative analysis of ablation structures and volumes.

While we our goal was to analyze commonly encountered SOPs, we did not analyze the presence of multiple onset patterns within the same seizure after the first five seconds (e.g., RSPT followed by LFVA). Quantitative analysis of SOPs may be helpful in refining categorization and should be addressed in future studies. While tools are in development to automate quantitative analysis, it is not routinely performed at many surgical centers. Contrary to quantitative measures, SOPs are readily clinically accessible and require no post-processing. Further efforts should be made to develop biomarkers utilizing this.

Although we found a subset of SOPs that predict SLAH failure, over half of patients with favorable SOPs still fail SLAH. One possibility is that refining the surgical approach to achieve complete ablation of mesial temporal structures may improve outcomes. Another possibility is that seizures may involve neocortical structures adjacent to hippocampus such as the parahippocampal gyrus and entorhinal cortex, or distant structures such as insular cortices or lateral neocortical temporal sites that were not sampled.^45^ We speculate that extending the SLAH targeting to include adjacent neocortical structures may improve outcomes for patients with favorable SOPs, but not for patients with unfavorable SOPs. Differentiating these scenarios, and determining how seizure network dynamics and SOPs are related, is an area in need of further study.

## 5. Conclusions

SOPs are useful in predicting two-year outcomes after sEEG-guided SLAH, consistent with prior literature in patients undergoing open resection. Repetitive spiking in the theta or alpha range and rhythmic slowing in the theta or delta range were strongly associated with a poor outcome.

Future studies are warranted to further understand the impact of less frequently encountered SOPs on seizure freedom, how integration of quantitative EEG and MRI analysis influence outcome prediction, and how seizure network dynamics underlie the emergence of specific seizure onset patterns.

## Data Availability

All data produced in the present study are available upon reasonable request to the authors

## Acknowledgements

Research in this manuscript was supported by the National Institute of Neurological Disorders and Stroke (NINDS) of the National Institute of Health (AJM – T32 NS07153; CAS – R01 NS084142, NS110669, and RF1-MH114276), S.T. has received support from the Department of Veterans Affairs, VISN1 Veterans Health Administration (V1CDA2022-68). K.A.D has received support from R01-NS-116504 and R01-NS-125137.

## References

1. Wiebe S, Blume WT, Girvin JP, Eliasziw M, Effectiveness, Efficiency of Surgery for Temporal Lobe Epilepsy Study G. A randomized, controlled trial of surgery for temporal-lobe epilepsy. N Engl J Med. 2001;345(5):311–8.

2. Youngerman BE, Save AV, McKhann GM. Magnetic Resonance Imaging-Guided Laser Interstitial Thermal Therapy for Epilepsy: Systematic Review of Technique, Indications, and Outcomes. Neurosurgery. 2020;86(4):E366–E82.

3. Drane DL, Willie JT, Pedersen NP, Qiu D, Voets NL, Millis SR, et al. Superior Verbal Memory Outcome After Stereotactic Laser Amygdalohippocampotomy. Front Neurol. 2021;12:779495.

4. Brotis AG, Giannis T, Paschalis T, Kapsalaki E, Dardiotis E, Fountas KN. A meta-analysis on potential modifiers of LITT efficacy for mesial temporal lobe epilepsy: Seizure-freedom seems to fade with time. Clin Neurol Neurosurg. 2021;205:106644.

5. Youngerman BE, Oh JY, Anbarasan D, Billakota S, Casadei CH, Corrigan EK, et al. Laser ablation is effective for temporal lobe epilepsy with and without mesial temporal sclerosis if hippocampal seizure onsets are localized by stereoelectroencephalography. Epilepsia. 2018;59(3):595–606.

6. Barot N, Batra K, Zhang J, Klem ML, Castellano J, Gonzalez-Martinez J, et al. Surgical outcomes between temporal, extratemporal epilepsies and hypothalamic hamartoma: systematic review and meta-analysis of MRI-guided laser interstitial thermal therapy for drug-resistant epilepsy. J Neurol Neurosurg Psychiatry. 2022;93(2):133–43.

7. Schevon CA, Weiss SA, McKhann G, Jr., Goodman RR, Yuste R, Emerson RG, et al. Evidence of an inhibitory restraint of seizure activity in humans. Nat Commun. 2012;3:1060.

8. Weiss SA, Banks GP, McKhann GM, Jr., Goodman RR, Emerson RG, Trevelyan AJ, et al. Ictal high frequency oscillations distinguish two types of seizure territories in humans. Brain. 2013;136(Pt 12):3796–808.

9. Smith EH, Schevon CA. Toward a Mechanistic Understanding of Epileptic Networks. Curr Neurol Neurosci Rep. 2016;16(11):97.

10. Eissa TL, Dijkstra K, Brune C, Emerson RG, van Putten M, Goodman RR, et al. Cross-scale effects of neural interactions during human neocortical seizure activity. Proc Natl Acad Sci U S A. 2017;114(40):10761–6.

11. Smith EH, Merricks EM, Liou JY, Casadei C, Melloni L, Thesen T, et al. Dual mechanisms of ictal high frequency oscillations in human rhythmic onset seizures. Sci Rep. 2020;10(1):19166.

12. Schiller Y, Cascino GD, Busacker NE, Sharbrough FW. Characterization and comparison of local onset and remote propagated electrographic seizures recorded with intracranial electrodes. Epilepsia. 1998;39(4):380–8.

13. Schuh LA, Henry TR, Ross DA, Smith BJ, Elisevich K, Drury I. Ictal spiking patterns recorded from temporal depth electrodes predict good outcome after anterior temporal lobectomy. Epilepsia. 2000;41(3):316–9.

14. Faught E, Kuzniecky RI, Hurst DC. Ictal EEG wave forms from epidural electrodes predictive of seizure control after temporal lobectomy. Electroencephalogr Clin Neurophysiol. 1992;83(4):229–35.

15. Gupta K, Cabaniss B, Kheder A, Gedela S, Koch P, Hewitt KC, et al. Stereotactic MRI-guided laser interstitial thermal therapy for extratemporal lobe epilepsy. Epilepsia. 2020;61(8):1723–34.

16. Zaher N, Urban A, Antony A, Plummer C, Bagic A, Richardson RM, et al. Ictal Onset Signatures Predict Favorable Outcomes of Laser Thermal Ablation for Mesial Temporal Lobe Epilepsy. Front Neurol. 2020;11:595454.

17. Malmgren K, Thom M. Hippocampal sclerosis--origins and imaging. Epilepsia. 2012;53 Suppl 4:19–33.

18. Spencer SS, Spencer DD. Entorhinal-hippocampal interactions in medial temporal lobe epilepsy. Epilepsia. 1994;35(4):721–7.

19. Dolezalova I, Brazdil M, Hermanova M, Horakova I, Rektor I, Kuba R. Intracranial EEG seizure onset patterns in unilateral temporal lobe epilepsy and their relationship to other variables. Clin Neurophysiol. 2013;124(6):1079–88.

20. Singh S, Sandy S, Wiebe S. Ictal onset on intracranial EEG: Do we know it when we see it? State of the evidence. Epilepsia. 2015;56(10):1629–38.

21. Jimenez-Jimenez D, Nekkare R, Flores L, Chatzidimou K, Bodi I, Honavar M, et al. Prognostic value of intracranial seizure onset patterns for surgical outcome of the treatment of epilepsy. Clin Neurophysiol. 2015;126(2):257–67.

22. Lagarde S, Buzori S, Trebuchon A, Carron R, Scavarda D, Milh M, et al. The repertoire of seizure onset patterns in human focal epilepsies: Determinants and prognostic values. Epilepsia. 2019;60(1):85–95.

23. Merricks EM, Smith EH, Emerson RG, Bateman LM, McKhann GM, Goodman RR, et al. Neuronal Firing and Waveform Alterations through Ictal Recruitment in Humans. J Neurosci. 2021;41(4):766–79.

24. Liou JY, Smith EH, Bateman LM, Bruce SL, McKhann GM, Goodman RR, et al. A model for focal seizure onset, propagation, evolution, and progression. Elife. 2020;9.

25. Spencer SS, Guimaraes P, Katz A, Kim J, Spencer D. Morphological patterns of seizures recorded intracranially. Epilepsia. 1992;33(3):537–45.

26. Wetjen NM, Marsh WR, Meyer FB, Cascino GD, So E, Britton JW, et al. Intracranial electroencephalography seizure onset patterns and surgical outcomes in nonlesional extratemporal epilepsy. J Neurosurg. 2009;110(6):1147–52.

27. Kane N, Acharya J, Benickzy S, Caboclo L, Finnigan S, Kaplan PW, et al. A revised glossary of terms most commonly used by clinical electroencephalographers and updated proposal for the report format of the EEG findings. Revision 2017. Clin Neurophysiol Pract. 2017;2:170–85.

28. Engel J, Jr. Update on surgical treatment of the epilepsies. Summary of the Second International Palm Desert Conference on the Surgical Treatment of the Epilepsies (1992). Neurology. 1993;43(8):1612–7.

29. van der Walt S, Colbert SC, Varoquaux G. The NumPy Array: A Structure for Efficient Numerical Computation. Comput Sci Eng. 2011;13(2):22–30.

30. Hunter JD. Matplotlib: A 2D Graphics Environment,. Comput Sci Eng. 9(3):90–5.

31. Davidson-Pilon C. Lifelines: survival analysis in Python. Journal of Open Source Software. 2019;4(40):1317.

32. Tao JX, Wu S, Lacy M, Rose S, Issa NP, Yang CW, et al. Stereotactic EEG-guided laser interstitial thermal therapy for mesial temporal lobe epilepsy. J Neurol Neurosurg Psychiatry. 2018;89(5):542–8.

33. Thorsteinsdottir J, Vollmar C, Tonn JC, Kreth FW, Noachtar S, Peraud A. Outcome after individualized stereoelectroencephalography (sEEG) implantation and navigated resection in patients with lesional and non-lesional focal epilepsy. J Neurol. 2019 Apr;266(4):910–20.

34. Sokolov E, Sisterson ND, Hussein H, Plummer C, Corson D, Antony AR, et al. Intracranial monitoring contributes to seizure freedom for temporal lobectomy patients with nonconcordant preoperative data. Epilepsia Open. 2022;7(1):36–45.

35. Park YD, Murro AM, King DW, Gallagher BB, Smith JR, Yaghmai F. The significance of ictal depth EEG patterns in patients with temporal lobe epilepsy. Electroencephalogr Clin Neurophysiol. 1996;99(5):412–5.

36. Alter AS, Dhamija R, McDonough TL, Shen S, McBrian DK, Mandel AM, et al. Ictal onset patterns of subdural intracranial electroencephalogram in children: How helpful for predicting epilepsy surgery outcome. Epilepsy Res. 2019 01;149:44–52.

37. Feng R, Farrukh Hameed NU, Hu J, Lang L, He J, Wu D, et al. Ictal stereo-electroencephalography onset patterns of mesial temporal lobe epilepsy and their clinical implications. Clin Neurophysiol. 2020;131(9):2079–85.

38. Grewal SS, Zimmerman RS, Worrell G, Brinkmann BH, Tatum WO, Crepeau AZ, et al. Laser ablation for mesial temporal epilepsy: a multi-site, single institutional series. J Neurosurg. 2018:1–8.

39. Donos C, Breier J, Friedman E, Rollo P, Johnson J, Moss L, et al. Laser ablation for mesial temporal lobe epilepsy: Surgical and cognitive outcomes with and without mesial temporal sclerosis. Epilepsia. 2018;59(7):1421–32.

40. Wu C, Jermakowicz WJ, Chakravorti S, Cajigas I, Sharan AD, Jagid JR, et al. Effects of surgical targeting in laser interstitial thermal therapy for mesial temporal lobe epilepsy: A multicenter study of 234 patients. Epilepsia. 2019;60(6):1171–83.

41. Satzer D, Tao JX, Warnke PC. Extent of parahippocampal ablation is associated with seizure freedom after laser amygdalohippocampotomy. J Neurosurg. 2021:1–10.

42. Liu DD, Lauro PM, Phillips RK, 3rd, Leary OP, Zheng B, Roth JL, et al. Two-trajectory laser amygdalohippocampotomy: Anatomic modeling and initial seizure outcomes. Epilepsia. 2021;62(10):2344–56.

43. Kang JY, Wu C, Tracy J, Lorenzo M, Evans J, Nei M, et al. Laser interstitial thermal therapy for medically intractable mesial temporal lobe epilepsy. Epilepsia. 2016;57(2):325–34.

44. Barkley AS, Sullivan LT, Gibson AW, Zalewski K, Mac Donald CL, Barber JK, et al. Acute Postoperative Seizures and Engel Class Outcome at 1 Year Postselective Laser Amygdalohippocampal Ablation for Mesial Temporal Lobe Epilepsy. Neurosurgery. 2022;91(2):347–54.

45. Barba C, Rheims S, Minotti L, Guenot M, Hoffmann D, Chabardes S, et al. Temporal plus epilepsy is a major determinant of temporal lobe surgery failures. Brain. 2016;139(Pt 2):444–51.

